# Safety and Efficacy of the combined use of ivermectin, dexamethasone, enoxaparin and aspirin against COVID 19

**DOI:** 10.1101/2020.09.10.20191619

**Authors:** Carvallo Héctor, Hirsch Roberto, Farinella María Eugenia

**Author notes:** NOTE: This Protocol and its trial were duly submitted to: http://ClinicalTrials.gov Identifier: NCT04425863 Eurnekian Public Hospital Protocol Record IDEA, Ivermectin, Dexametasone, Enoxaparin and Aspirin as treatment for Covid 19*.

## Abstract

From the first outbreak in Wuhan (China) in December 2019, until today the number of deaths worldwide due to the coronavirus pandemic exceeds eight hundred thousand people and the number of infected people arises to more than 25 million.

No treatment tested worldwide has shown unquestionable efficacy in the fight against COVID 19, according to NICE reports.

We have designed an experimental treatment called IDEA based on four affordable drugs already available on the market in Argentina, based on the following rationale:

- **I**vermectin solution at a relatively high dose to lower the viral load in all stages of COVID 19
- **D**examethasone 4-mg injection, as anti-inflammatory drug to treat hyperinflammatory reaction to COVID-infection
- **E**noxaparin injection as anticoagulant to treat hypercoagulation in severe cases.
- **A**spirin 250-mg tablets to prevent hypercoagulation in mild and moderate cases

Except for Ivermection oral solution, which was used in a higher dose than approved for parasitosis, all other drugs were used in the already approved dose and indication. Regarding Ivermectin safety, several oral studies have shown it to be safe even when used at daily doses much higher than those approved already.

A clinical study has been conducted on COVID-19 patients at Eurnekian Hospital in the Province of Buenos Aires, Argentina. The study protocol and its final outcomes are described in this article. Results were compared with published data and data from patients admitted to the hospital receiving other treatments.

None of the patient presenting mild symptoms needed to be hospitalized. Only one patient died (0.59 % of all included patients vs. 2.1 % overall mortality for the disease in Argentina today; 3.1 % of hospitalized patients vs. 26.8 % mortality in published data). IDEA protocol appears to be a useful alternative to prevent disease progression of COVID-19 when applied to mild cases and to decrease mortality in patients at all stages of the disease with a favorable risk-benefit ratio.

## Introduction

In late December 2019, the incidence of atypical pneumonia cases of unknown cause was reported in the Chinese city of Wuhan. PCR (Polymerase Chain Reaction) studies found a new coronavirus, named SARS-CoV-2. The disease caused by the virus has been named COVID-19.

To design a treatment protocol using affordable, already marketed drugs, we have considered the information already available about SARS-CoV-2 and COVID-19.

SARS-CoV 2 proved to be remarkably similar to SARS-CoV, the only significant different bein a furin-binding domain in the SARS-CoV-2 protein S, which may expand tropism or increase virus transmission (1). Studies on several RNA viruses have revealed a potential role for IMPα / β1 during infection (2). Moreover, entry of SARS-CoV-2 into cells is facilitated by its optimized binding to ACE2 (angiotensin conversion enzyme 2) (3). This enzyme acts a receptor for SARS-CoV-2 and is found in multiple tissues (4, 5, 6), including alveolar epithelium of the lung, arterial and venous endothelium, smooth muscle, renal tubular epithelium, oropharyngeal mucosa and epithelium of the small intestine, largely explaining the clinical presentation of patients with COVID-19. Ivermectin, an antiparasitic drug, already known to inhibit RNA viruses by interfering with IMP IMPα / β1 (2 7prima) has been shown to inhibit SARS-CoV-2 in vitro (8). Ivermectin is already available as oral tablets and drops on the market in several countries and may be useful to treat COVID-19. Therefore, we have included Ivermectin oral solution as an antiviral drug in our treatment protocol. We selected an oral dose much higher than that being used for parasitosis but within safety margins according to previous clinical trials published in the literature (9, 10). Guzzo et al. (9) administered up to three doses of 60 mg to adults and Levy reports doses higher than 500 µg/kg in children and 400 µg/kg in children below 5 years of age with good safety profile. IDEA protocol includes doses of 24, 36 and 48 mg on days 0 and 7 of treatment equivalent to doses of ca. 300, 450 and 600 g/kg for mild, moderate and severe cases of 80-kg adults, which are lower than the maximum doses already tested in adult humans. The reason of this decision is that,in spite of pharmacokinetic data available show that the maximum plasma concentration are far below the concentration needed to achieve inhibition of SARS-CoV-2 in-vitro (11), we consider ivermectin to be potentially effective to fight COVID-19 because it tends to be distributed in different organs due to its lipophilicity and could reach higher levels in high ACE2 expressing organs than in bloodstream (12). Posology consisted of two doses of Ivermectin on day 0 (start of treatment) and 7. This time interval avoids accumulation of drug according to plasma half life reported in literature (9, 10). Ivermectin oral solution was used because previous studies allowed the hypothesis of a better bioavailability (higher plasma concentrations) (13).

A significant percentage of patients, ca. 30 – 50 % of total, suffering from COVID-19 may not present any symptoms (14). Most patients develop mild symptoms. However, a substantial percentage of patients develop moderate to severe forms of the disease, needing hospital care and even ICU treatment. Mortality rate is approximately 26.8 % in these patients (15).

The most frequent symptoms of COVID-19 are fever, cough, dyspnea, myalgias or fatigue. Other reported symptoms are bilateral conjunctival injection without associated secretions, hypogeusia, skin rash and hyposmia (16). A group of patients with severe forms of COVID-19 develop a cytokine storm syndrome (16). Several authors link this cytokine storm syndrome to secondary hemophagocytic lymphohistiocytosis (abridged sHLH), a poorly recognized hyperinflammatory syndrome leading to multiple organ failure and death, triggered by viral infections (17, 18). The main features of sHLH include unremitting fever, cytopenias, and hyperferritinaemia. Pulmonary involvement is present in approximately 50% of patients. A cytokine profile that resembles sHLH is associated with the severity of COVID-19 disease (19). Mortality predictors from a recent multicenter retrospective study of 150 confirmed cases of COVID-19 in Wuhan, China included elevated ferritin (mean 1297.6 ng / ml in non-survivors versus 614.0 ng / ml in survivors; p < 0.001) and IL-6 (p < 0.0001), suggesting that mortality could be due to viral hyperinflammation(19). Therefore, we have included Dexamethasone injection in our treatment protocol for moderate and severe cases to treat hyperinflammation (20).

Even though COVID-19 is primarily a respiratory disease, accumulating data suggests it to be profoundly prothrombotic (21, 22, 23, 24, 25, 26, 27, 28). In fact, microthrombosis in different locations has been repeatedly reported in patients with COVID 19. This symptom is due to a hypercoagulable state (21, 22, 24) caused by this disease. Therefore, we have included aspirin tablets as antithrombotic agent for mild and moderate cases (29) and enoxaparin injection as anticoagulant for severe cases in IDEA treatment protocol (30, 31, 32).

Thus, this four-drug treatment protocol may allow to control viral load, as well as the most serious symptoms leading patients to ICU treatment and death. In moderate to severe cases we added ventilation and standard supportive care. We present in this article the final outcomes of a clinical trial on COVID-19 patients treated with this protocol.

## Materials and Methods

### Materials

Ivermectin 0.6 mg/ml solution, Dexamethasone 4-mg injection, Enoxaparin injection and Aspirin 250-mg tablets purchased from the Argentinean market were used.

### Study design

The study was a single-center, prospective clinical trial run at Eurnekian Hospital in the Province of Buenos Aires, Argentina from May 2020 to July 2020. For ethical reasons, all patients received treatment and supportive care.

### Patients

Patients included in this study protocol were male and female persons not less than 5 years old, with a positive rt-PCR diagnosis of COVID-19 performed on nasal swab specimens, able to provide informed consent and not participating in any other clinical study. polymerase chain reaction (PCR), no participation in other clinical trials during the study period, and able to provide informed consent. Pregnant women and persons with previous reports of allergy to any of the drugs included in the treatment were excluded.

### Treatment

Symptoms were classified as mild and severe according to the following table:

| MILD SYMPTOMS | SEVERE SYMPTOMS |
| --- | --- |
| Fever not above 38.5 °C<br>Isolated diarrheal episodes<br>Hyposmia or Hypogeusia<br>Mild desaturation (93 – 96 %)<br>Dyspnea without matter<br>Polymyoarthralgias, Persistent headache,<br>Abdominal pain | Fever above 38.5 °C<br>Diarrhea (more than 3 daily depositions)<br>Flictenular conjunctivitis<br>Strong desaturation (92% or less)<br>Tachypnea (FR> 25 / minute) |

Disease stages were classified according to the following table:

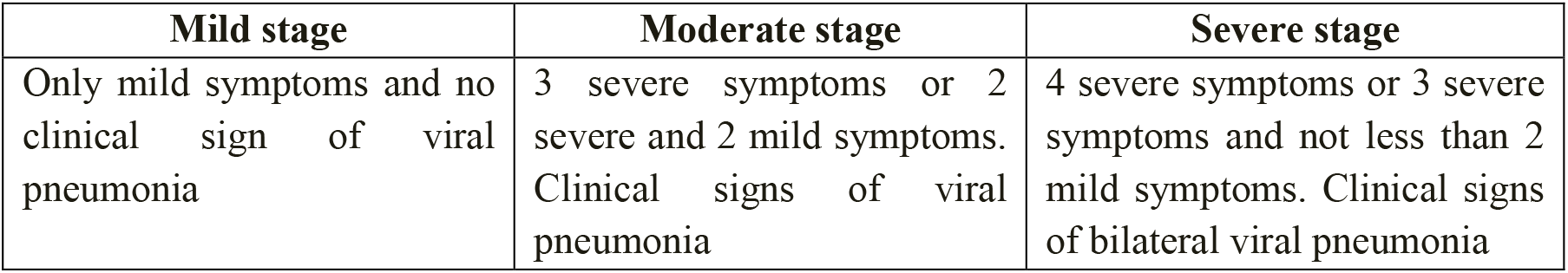

| Mild stage | Moderate stage | Severe stage |
| --- | --- | --- |
| Only mild symptoms and no clinical sign of viral pneumonia | 3 severe symptoms or 2 severe and 2 mild symptoms. Clinical signs of viral pneumonia | 4 severe symptoms or 3 severe symptoms and not less than 2 mild symptoms. Clinical signs of bilateral viral pneumonia |

The following treatmen protocol was used on each case according to the following table:

The following treatment protocol was used on each case according to the following table:
| **DISEASE SEVERITY** | **IVERMECTIN ORAL SOLUTION** | **DEXAMETHASONE** | **ANTITHROMBOTIC /ANTICOAGULANT** | **VENTILATION** |
| --- | --- | --- | --- | --- |
| **Mild stage** | 24 mg on days 0 and 7 | None | Aspirin 250-mg tablet once daily for at least 30 days | No |
| **Moderate stage** | 36 mg on days 0 and 7 | Dexamethasone 4- mg injection daily | Aspirin 250-mg tablet once daily for at least 30 days | Low Flow Washed Oxygen or Oxygen Concentrator |
| **Severe stage** | 48 mg via gastric cannulae on days 0 and 7 | Dexamethasone 4- mg injection on day daily. | Enoxaparin 100 UI/kg (ca. 1 mg/kg) daily | Mechanical Ventilation |

Patients at mild stage of COVID-19 were treated as outpatients. They came to the hospital to receive drugs and remote follow-up via mobile phone was implemented. Patients at moderate stage of disease were immediately admitted to ward care. Patients at severe stage of COVID-19 were immediately admitted to ICU.

The day on which each patient starts treatment is numbered as “day 0”. All other days are numbered in relation to this one.

Inpatients were discharged from ICU to hospital ward care when their symptoms were compatible with moderate stage disease.

Inpatients were discharged home from hospital when they tested negative for COVID-19 as determined by rt-PCR on specimens obtained from nasal swabbing or when their symptoms were compatible with mild stage of disease.

Outpatients were considered cured after one negative COVID-19 determination by rt-PCR on specimens or 10 days without any symptoms.

### Ethics Committee aproval

The study was approved both by the ethics committee of the hospital and the County Ethics Committee, in accordance with the Declaration of Helsinski and its ammendments. All included patients signed an informed consent before inclusion. This study was registered in http://ClinicalTrials.gov website under the identifier number: NCT04425863.

### Outcomes

The primary outcomes were:

- Percentage of patients progressing from mild to moderate or severe stages of disease
- Mortality rate by day 30

The secondary outcomes were safety outcomes related to treatment adverse events and dose adjustments for any of the drugs used.

### Statistical analysis

Data were collected and recorded in MS-Excel spreadsheets and processed. Demographic data, prevalence of different comorbidities and outcomes were calculated. The outcomes of the study were compared with data from the literature and, in the case of moderate to severe cases, with a group of patients admitted to the hospital in the same period of time who did not join the study protocol and received other treatments.

## Results

### Population characteristics

A total of 167 patients were included. All of them had confirmed COVID19 infection by the rtPCR method. Average age was 55.7 years, 48.5 % were female and 51.5 % were male. The stages of disease of all included patients when they join the study were:

| **Disease stage at inclusion** | **Patients (n)** | **Average age (years)** | **Sex (female/male %)** |
| --- | --- | --- | --- |
| Mild | 135 | 55.7 | 48.5 / 51.5 |
| Moderate to severe | 32 | 59.7 | 57.5 / 42.5 |
| Total | 167 | 56.5 | 50,2 / 49,8 |

From the moderate to severe cases included, 23 (71.9 %) presented at list one risk factor.

### Disease progression and mortality

All the135 patients who joined the study at a mild stage of COVID-19 did not worsen illness and had no need of hospitalization of any kind.

Regarding the remaining 32 patients, only one of them died. This patient had been included already at a severe stage of disease. The remaining 31 patients did not worsen during treatment.

### Comparison with other treatments

Overall mortality rate of patients treated according to IDEA protocol was 0.59 % (1 death in 167 treated cases). As a comparison, estimated overall mortality rate in Argentina is approximately 2.1 % (official data by September 2^nd^, 2020).

Regarding moderate to severe cases, i.e. patients needing hospitalization, only 1 patient out of 32 receiving IDEA treatment died (3.1 %), whereas mortality rate published in articles from Spain, Italy and Spain is ca. 25 %. Moreover, a group of 12 patients were hospitalized in Eurnekian hospital in the same period but did not receive IDEA treatment. Three of them died, thus presenting a mortality rate of 25 %, i.e. significantly higher than that of those receiving IDEA treatment.

### Adverse events

Only one patient suffered from a serious adverse event. It was a patient who had previous history of gastric ulcer and contracted one during treatment, probably caused by Dexamethasone injection. The problem was immediately solved with iced water through oropharyngeal cannulae and omeprazole, without discontinuation of treatment or dose adjustment.

### Dose adjustments

No dose adjustment was necessary.

## Discussion

IDEA treatment has proved to be efficacious in preventing worsening of the symptoms of COVID-19 in practically all treated patients.

Overall mortality rate in patients treated according to IDEA protocol is significantly lower (0.59 %) than that of the infected population in Argentina (2.1 % by September 2^nd^, 2020) and other countries, like Brazil (3.1 % by September 2^nd^, 2020) today. Quite strikingly, only 1 out of 32 hospitalized patients died of COVID-19 when treated according to the IDEA protocol, whereas the published data show a 26.8 % mortality for inpatients (15), i.e. 8 deaths instead of 1. A similar mortality rate of 25 % (3 deaths) was observed in a group of 12 patients admitted to Eurnekian hospital in the same period, who received other treatments.

Regarding disease progression, no patient with mild COVID-19 progressed to moderate or severe disease after treatment according to IDEA protocol. Moreover, even though we did not have a quantitative follow-up of time to absence of symptoms, we have observed it to be less than 3 days in many mild cases.

We consider that our data, even though not being placebo-controlled due to ethical reasons, allow us to conclude that IDEA protocol may be of use to help stop COVID-19 progression and reduce hospitalization and mortality.

Based on the outcomes of this study, a possible preventive strategy for COVID-19 in communities of high viral circulation might consist of an oral dose of ivermectin lower than 24 mg (proposed 12 mg) regularly administered once a week (approximately one incubation period) to low-risk people for a limited period of time, while high risk population remains isolated. This dose might be enough to reduce viral load at a low level to keep COVID-19 at a mild stage, without eliminating SARS-CoV-2 completely, so that immunity against SARS-CoV-2 is developed individually to finally reach herd immunity (“immunizing effect”). This hypothesis is worth further exploration for the prevention of transmission in healthcare workers and close contacts, and, if successful, may be further applied for prevention in the community. In the present situation in some American countries like Brazil and Argentina, this could help reduce overall mortality in the absence of a vaccine.

## Conclusions

Given the rapid advance of COVID-19 pandemic in Argentina and several other countries and the lack of evidence of efficacy of any other treatments, we consider IDEA protocol to be a scientifically based, low-cost treatment and sustainable healthcare strategy with a favorable benefit-risk ratio, based on the results reported in this article: reduction in overall mortality to one fourth, reduction in mortality of hospitalized patients to one eighth and no progression of mild cases to moderate or severe cases. It should be emphasized that the earlier the treatment starts, the better. Mild cases early treated according to our protocol did not progress to more severe stages. Furthermore, even though not exactly quantitated, we noticed the best results in the treatment of mild cases, most of them recovering in less than 3 days. The application of early treatment is consistent with the principles of medicine: even the most efficient therapeutic measures will lose efficacy if they are applied too late. In conclusion, IDEA seems to be an adequate treatment strategy for pandemic COVID-19 disease at all stages of disease, but specially so, if applied at the earliest possible stage thereof.

## Data Availability

Data available on request.

